# Voices from the front line: community health workers’ perspectives on effectiveness and needs in Madagascar’s tuberculosis response

**DOI:** 10.64898/2025.12.23.25342944

**Authors:** Onja Gabrielle Ravololohanitra, Alphonsine Razanamiarana, Michel Tiaray Harison, Justine Nafeno, Lydia Raveloarisoa, Harizaka Emmanuel Andriamasy, Haingotiana Razafimanantsoa, Anne Neumann, Nadine Muller

## Abstract

**Background:** Community health workers (CHWs) are central to primary health care delivery in Madagascar, yet their perspectives on what enables effective work are rarely centered in programme design and implementation. Evidence is needed on the support CHWs receive in practice and the priorities they identify to strengthen effectiveness and equity.

**Methods:** We conducted a qualitative study nested within the formative phase of a participatory action research initiative aimed at strengthening Madagascar’s community health workforce. Tuberculosis (TB) programme support was used as a tracer condition for universal health coverage-relevant service delivery. Five focus group discussions were held with 26 CHWs in August-September 2024 in two contrasting regions (Analamanga and Atsimo-Andrefana). Discussions were conducted in Malagasy and its dialects, audio-recorded, transcribed, translated into French, and analyzed using inductive reflexive thematic analysis.

**Results:** CHWs articulated five interlinked priorities for effectiveness in the TB response: (1) stable and equitable socio-economic protection; (2) standardized, continuous, and practical training; (3) formative and collaborative supervision; (4) reliable provision of tools, equipment, transport, and personal protective equipment; and (5) recognition and integration within communities and the health system. CHWs emphasised reliability and fulfilment of promised entitlements and described fragmentation across implementing partners as a key source of inequity, uncertainty, and demotivation.

**Conclusions:** CHWs’ accounts point to a persistent implementation gap between policy commitments and day-to-day support. Strengthening CHW effectiveness in Madagascar will require coordinated national stewardship and partner accountability to deliver a predictable minimum support package and to include CHWs meaningfully in programme design and evaluation.

## Introduction

Community health workers (CHWs) provide essential primary health care in many low- and middle-income countries, particularly across sub-Saharan Africa. Their close ties to communities, socially and geographically, support health promotion, early disease detection, adherence support, timely referral, and long-term follow-up, thereby improving the availability, accessibility, acceptability, and quality of health services [1].

Global momentum increasingly recognises CHWs not only as programmatic assets but also as rights-holders whose work must be protected. The 2018 Declaration of Astana [2] and WHO’s guideline on optimising CHW programmes [3] position CHWs at the centre of primary health care and specify obligations for fair remuneration, training, supportive supervision, and adequate supplies. Evidence from Brazil, Ethiopia, and Nepal [4] suggests that when CHWs are integrated with clear roles, equitable pay, and non-punitive supervision, progress towards universal health coverage (UHC) accelerates [5]. The 2023 Monrovia Call to Action urges governments and partners to fully fund, integrate, and safeguard CHWs [6]. Yet despite this momentum - and the growing visibility of advocacy platforms that amplify CHW voices – CHWs’ participation in decision-making and programme design often remains limited [7]. Consequently, in many settings, CHWs continue to face precarious or opaque compensation, uneven access to training and tools, limited opportunities for participation and accountability, and weak recognition within decision-making structures [8–10].

Madagascar, an island nation of about 32 million people [11], faces substantial primary health-care delivery challenges, with a UHC service coverage index of 35 out of 100 in 2019 [12]. Nearly 59% of the population lives in rural areas [13], where CHWs play an essential role in extending access to care and advancing progress towards UHC. CHWs are officially recognised in national policy, including the *Politique Nationale de Santé Communautaire* (PNSC), and largely operate on a voluntary basis [14]. Programme implementation involves multiple partners, contributing to fragmentation across health topics and gaps in accountability. In this plural landscape, CHWs may be uncertain about their entitlements (e.g., compensation, training, equipment, transport), and the CHW workforce is seldom systematically involved in programme design and evaluation. As a result, CHWs’ own accounts of what strengthens (or constrains) their effectiveness are rarely centered, limiting evidence on what they receive in practice and whether support programmes adequately enable them to work effectively.

Using tuberculosis (TB) programme support as a tracer condition to UHC, this qualitative study foregrounds CHWs’ perspectives from two contrasting regions and elicits their recommendations for improving CHW effectiveness in the TB response.

## Methods

### Study design

We conducted a qualitative study nested within the formative phase of a participatory, intersectoral action research initiative aimed at strengthening Madagascar’s community health workforce [15]. CHWs were engaged as partners in identifying challenges, generating evidence, interpreting findings to inform practice-oriented improvements. To anchor the analysis in specific service delivery realities while examining broader system enablers and barriers, we used TB as a tracer condition. Madagascar’s persistently high TB burden (estimated incidence of 233 cases per 100,000 population, unchanged since 2013) makes TB a sensitive indicator of service acceptability, accessibility, and quality, including stigma-aware communication, safe working conditions, and reliable referral and long-term follow-up [16].

### Study setting

Madagascar, an island nation in the Indian Ocean. An estimated 80% of the population lived below the international poverty line (<US$2.15/day, 2017 purchasing power parity) between 2022-2024 [17]. The economy is based mainly on subsistence farming, livestock, fishing, and the informal sector. The health system is organised as a pyramid with Basic Health Centres (Centres de Santé de Base - CSB) at community level, district hospitals, and regional or central referral hospitals. A national network of approximately 50,700 volunteer CHWs serves on the front line [18], providing prevention, screening, and patient support, and linking communities to CSBs. National policy documents outline the overarching context and working conditions, such as the national strategic plan for strengthening community health [14] and the Community Activities Package guide (Guide PAC) [19].

This qualitative study was conducted in two epidemiologically, socioeconomically, and geographically contrasting regions. Analamanga, which includes the capital Antananarivo, is largely urban and peri-urban, with high population density, relatively higher service coverage, and a TB notification rate of 176 per 100,000 in 2024 [20]. Atsimo-Andrefana, the country’s largest administrative region in the southwest, is predominantly rural, with low population density, sparse health infrastructure, and high multidimensional poverty [12]. The TB notification rate in this region was 313 per 100,000 in 2024 [20]. Access to care in this region is largely constrained by distance, transport, and household economic barriers [21]. Including CHWs from both regions was a purposive decision to capture the diversity of operating conditions and to identify equity gaps that shape CHWs’ ability to engage in TB-related services.

### Sampling and selection of participants

Participants were recruited using a two-stage purposive sampling strategy. In the first stage, CSBs were selected in each region based on (i) TB activity during the preceding 12 months, (ii) catchment profile (urban, peri-urban, or rural), and (iii) logistical accessibility. In the second stage, CHWs from the selected CSBs were invited to participate if they were (1) aged 18 years or older, (2) currently active in their role, and (3) engaged in the TB response within the past 12 months. Recruitment and discussion-group composition intentionally balanced gender, tenure, role, and catchment profile to capture a wealth of perspectives and field realities. Participants were recruited between 19/08/2024 and 25/09/2024, during which time five focus group discussions (FGDs) were conducted.

### Data collection

FGDs were conducted in Malagasy and its respective dialects - Merina (highlands/Antananarivo), Mahafaly, Antandroy, and Masinkoro (Atsimo-Andrefana) - by a trained qualitative research team (coordinator, lead moderator, note taker). The data collection team had no prior relationship with participants. All moderators and note-takers were external to the communities and were not affiliated with local health facilities or community programs, reducing the risk of social desirability bias linked to familiarity. A local partner organization assisted with logistical arrangements and participant outreach, but did not take part in data collection or analysis.

The sessions lasted an average of 88 minutes. We used a semi-structured, pilot-tested thematic guide to explore CHW recruitment, roles, working conditions, initial and refresher training, financial and non-financial incentives, supervision practices, professional identity, and factors influencing overall and TB-specific commitment. Participants were asked to propose concrete recommendations for improvements in three areas: (i) effectiveness in current roles, (ii) working conditions, and (iii) additional support needs. Each discussion concluded with a co-design segment to generate recommendations.

With written and oral informed consent, all discussions were audio-recorded, transcribed verbatim, and translated into French under bilingual team supervision to preserve meaning and cultural nuance. Discrepancies were resolved by consensus. Participants who travelled to attend the discussions received reimbursement for transportation expenses in line with Malagasy research practice.

### Data analysis

The French transcripts were analyzed using an inductive, reflexive thematic approach within a constructivist-interpretivist orientation. The analysis was iterative, beginning with several close readings of the transcripts to familiarize the research team with the content, identify recurring ideas, and gradually develop a coding framework. As the analysis progressed, a dynamic codebook was created. Text segments were manually coded according to initial categories, which were refined as new codes. These were renamed, merged, split, or adjusted as understanding deepened [22,23]. This flexibility allowed the categories to capture the diversity and nuance of participants’ accounts.

The analysis was carried out by the first author, who managed the entire coding and interpretation process. At key stages, particularly for potentially ambiguous or complex segments, the analyst returned to the original-language transcripts and audio recordings to clarify meaning before finalizing codes. A second researcher was also involved to review specific excerpts and discuss the relevance or formulation of certain codes.

The organization of codes, the identification of relationships among them, and the final structuring into themes and subthemes were conducted using NVivo software (QSR International). Themes were defined based on their internal coherence, relevance to the research question, and grounding in participants’ narratives. Reporting follows the Consolidated Criteria for Reporting Qualitative Research (COREQ) checklist (**S1 Checklist**).

### Ethics and data protection

The study was approved by the Comité d’évaluation éthique pour la recherche biomédicale (CEER) at the National Public Health Institute (Institut National de la Santé Publique et Communautaire, INSPC) in Madagascar (003/2023_CEER INSPC/IRB) and by the Ethics Committee of Charité - Universitätsmedizin Berlin in Germany (EA2/083/23). Oral and written informed consent were obtained from all participants. Anonymity was maintained at all stages of the research, and all data were de-identified prior to translation. Audio recordings and transcripts were anonymized and assigned unique codes before being securely stored in a restricted-access drive, accessible only to members of the research team.

## Results

A total of 26 CHWs participated in five FGDs: three in the Analamanga region and two in the Atsimo-Andrefana region. The participants’ characteristics are presented in **Table 1**.

**Table 1.** Participant characteristics for CHW focus group discussions, Madagascar, 2024. CHW = community health worker, TB = tuberculosis.

| Characteristic | Category | Overall<br>n=26 (%) | Analamanga<br>n=14 | Atsimo-Andrefana<br>n=12 |
| --- | --- | --- | --- | --- |
| **Gender** | Female | 16 (62.5) | 13 | 3 |
|  | Male | 10 (37.5) | 1 | 9 |
| **Age (years)** | Range | 26 – 74 | 38 - 74 | 26 – 62 |
| **CHW role** | Generalist | 15 (57.7) | 6 | 9 |
|  | TB-focused | 11 (42.3) | 8 | 3 |
| **Area** | Urban | 10 (38.5) | 10 | 0 |
|  | Peri-urban | 3 (11.5) | 3 | 0 |
|  | Rural | 13 (50.0) | 0 | 13 |

The analysis generated five interlinked themes describing CHWs perceived needs to improve their effectiveness in Madagascar’s TB response: (1) stable and equitable socio-economic protection; (2) standardized, continuous training; (3) formative and collaborative supervision; (4) reliable tools, equipment, and transport; (5) and recognition and integration within the health system.

### Stable and equitable socio-economic protection

CHWs consistently described the need for comprehensive and stable socio-economic protection to enable and sustain their engagement. They reported a dual source of frustration: inadequate compensation and a perceived failure to uphold promised terms:

> “There is a small fee for completing this report (monthly TB case activity report). You will receive your payment in a month. And the work is done, but the low pay doesn’t match what was promised (by the program or supervisors).” (male CHW, rural Atsimo-Andrefana)This testimony highlights a gap between expectations and reality and a perceived breach of the moral contract between CHWs and partner institutions. Such experiences were described as profoundly undermining motivation and trust and reinforcing a sense of injustice.

CHWs also expressed a need for a basic level of economic security, enabling them to carry out their role with dignity, as well as formal recognition commensurate with their commitment. For several, the establishment of a professional status, including a regular salary and basic social protections, was deemed essential for long-term commitment. Others emphasized fixed, regular, and transparent compensation, whether as a salary, stipend, or allowance, as a matter of social justice, recognition, and minimum economic stability:

> “If we were given a small compensation of 100,000 Ar (22.5$) per month, it would be a way to make this task more acceptable to us, provided we had 100,000 Ar per month. This would be a change to implement.” (male CHW, rural Atsimo-Andrefana)

The harmonized implementation guide for community health programs in Madagascar [14] provides for financial incentives for CHWs. However, failure to adhere to these provisions has been perceived as contributing to significant disparities in remuneration, which are considered arbitrary and demotivating. Within the same district, some CHWs have received one-off allowances through NGO-led projects, while others, equally active, remain unpaid. This reflects the fragmentation of the community health system, in which NGOs operate independently with varying priorities and remuneration policies, often poorly aligned with national guidelines. As a result, many CHWs work for multiple NGOs simultaneously, without coordination or consistent social benefits. The lack of transparency and fairness was seen to foster a strong sense of injustice:

> “(…) We work as CHWs to accomplish tasks, and you (the NGOs) say that you (CHWs) will be helped by things like a telephone to communicate between us. So, for example, there are eight of us. And then only two of them get these things, even though they’ve all done the same job, which creates demotivation among us. The others get nothing, so it’s just two people who benefit.” (male CHW, rural Atsimo-Andrefana)

Unclear or perceived unfair distribution of benefits were described to undermine cohesion between CHWs and weaken collective commitment. Participants recommended improved payment organization and transparency, including a clear display of amounts due, predictable schedules, and verification mechanisms.

Many also recommended performance-based incentives based on criteria they considered objective (such as number of TB cases referred, quality of patient follow-up, engagement in sensitization), while acknowledging that some indicators are harder to measure.

Finally, CHWs stress the need for social support in the event of illness or serious personal events (e.g., accidents, pregnancy, family death):

> “When we’re sick, we go to the CSB2 (community health center) to get medicine. We’ve already spoken directly to the general manager, but he replied that it was voluntary work and that he couldn’t help us because it would become too much if they had to give us medicines. But sometimes we really need this little support, even for basic medicines, because sometimes we don’t have the money to buy it, given that we do this work voluntarily and our income isn’t even enough to buy medicines.” (female CHW, urban Analamanga).

This highlights a key weakness of the volunteer-based model in the given resource-limited setting: the absence of any safety net for CHWs, even to cover their personal basic health needs.

### Standardized, reality-grounded training

Training was perceived as a key lever to enhance effectiveness in the TB response. Many emphasized the need for ongoing, structured, and relevant training covering both technical knowledge and practical field skills. Regular refresher sessions were requested on TB-specific topics such as symptom detection, case management, adherence monitoring, and referral procedures. A single initial training was viewed as insufficient given evolving practices and operational challenges:

> “The training we receive is standard and very effective for us, but it needs to be reinforced. The last training session took place three years ago, and there should be new sessions to strengthen our capacity.” (male CHW, rural Atsimo-Andrefana)

CHWs called for training that includes transversal and relational skills such as communication, handling resistance, and adapting messages to vulnerable groups given the role of stigma and fear in hindering case detection and treatment adherence:

> “We have to go through training before becoming CHWs, because we never know what the people around us might pass on to us. If we don’t receive training—especially in terms of health—we won’t know how to handle it. We may be good, but not as good as we think. That’s why training is essential.” (female CHW, peri-urban Analamanga)

Another recurrent concern was perceived inequity in access to training, with opportunities varying widely by location and implementation partners, disfavouring specifically CHWs in rural and remote areas:

> “In our area, we’ve never had any refresher training. But we hear that in other communities, they have received new training on tuberculosis. Why not us?” (male CHW, rural Atsimo-Andrefana)

Participants therefore insisted on nationally standardized training to ensure a shared base of knowledge and skills, tailored to local realities, such as limited mobility, poor digital access, and cultural differences in rural areas that request adaptation of content and teaching methods:

> “Our suggestion is that you (the system)watch over us and support us. We frequently need training to strengthen our capacity and ensure that our knowledge is comprehensive.” (female CHW, rural Atsimo-Andrefana)

They also stress that training must be practical and grounded in reality, such as the use of daily reporting tools, field communication techniques adapted to the local context, concrete problem-solving, and simulated home visits.

### Formative and collaborative supervision

Many CHWs perceived the current supervision model as top-down or punitive and disconnected from the realities of their work. They call for a shift from a control-based approaches to formative, supportive, and collaborative supervision focused on guidance rather than fault-finding and punishment:

> “….as soon as you leave at 8 a.m. for the appointment in the fokontany, there is already a supervisor waiting for you. They accompany you everywhere, tirelessly, to the households of the target population. And from WHO as well — around noon, they called us saying: ‘We have arrived to inspect your work.’ Then we visited all the households. We checked the child’s finger to see whether they had received the vaccine or not, and whether they had taken the medication or not. We asked the parents these questions and took photos of the child’s finger.” (female CHW, urban Analamanga)

Participants specifically suggest regular, structured supervision based on dialogue, mutual respect and encouragement. A supportive approach – described as dynamic, motivating, and positive - was preferred:

> “Supervision shouldn’t be frightening; it should enable us to get on with the job and move it forward. It’s not there to scare us, but to ensure that we get the job done.” (male CHW, rural Atsimo-Andrefana)

CHWs recommended simple, accessible tools adapted to their profiles (often low literacy levels, limited connectivity) to help monitor activities, track progress, and identify training needs - without adding unnecessary workload. Supervision was also seen as an extension of training that reinforces skills, adjust practices, and flags areas for further learning:

> “Supervision isn’t a bad thing; it’s beneficial. It’s what allows you to see the work that’s not yet finished or that needs to be continued Supervision allows you to understand your own work, to see where there are little mistakes and correct them, and to take your work further. It makes you evolve, you say to yourself, “ah, is this what it means?” (male CHW, rural Atsimo-Andrefana)

Several CHWs also proposed involving local leaders in certain aspects of supervision to strengthen community and improve local coordination and expectations - especially in sensitive areas like TB screening:

> “If the chef Fokontany (local village lead) knows what we’re doing, he can support us. Otherwise, people ask us: who sent you? Why are you coming to my house?” (female CHW, urban Analamanga)

Given the elevated stigma surrounding TB, participants noted that home visits, presumptive case investigations, and referrals can be poorly perceived by families, giving rise to resistance or fear:

> “Convincing the patient to agree to an analysis is really difficult. As my colleague said, for us Madagascans it’s a shame to have the disease, and convincing them is still difficult, even to bring them here.” (female CHW, urban Analamanga)

Locally rooted supervision would help to overcome these misunderstandings by consolidating the social and institutional acceptability of CHWs’ work.

### Access to work tools, equipment, and transport

A frequently cited obstacle was disrupted availability of essential tools and resources for carrying out duties. These prerequisites were viewed as needing systematic and permanent rather than exceptional provision:

> “We CHWs, since we’re now talking about tuberculosis, should have a specific uniform as promoters, because ours, the old one, is in a bad shape, it’s too old! It hasn’t been replaced in nearly 20 years, we’ve only had one since the beginning, and what’s more, it’s bad. For example, we work in summer! We need raincoats, and we also need something to carry the equipment we use to raise awareness about tuberculosis! So, we need bags.” (female CHW, peri-urban Analamanga)

The needs expressed fell into two main categories: Basic functional equipment and tools to improve feasibility and efficiency.

#### Basic functional equipment

This includes forms, registers, and health education materials (e.g., posters, brochures). Their absence led CHWs to improvise or use personal resources, which participants felt affecting quality, consistency, and credibility:

> “For materials too, when we go out into the field, they should be complete. You (the system) should also give materials to the CHWs to help them explain to the people they are sensitizing.” (CHW, female, peri urban area, Analamanga).

#### Tools to improve feasibility and efficiency

CHWs reported often walking for hours to reach patients or health centers – a physically demanding and financially costly effort. Without access to bicycles or travel allowances (called “solom-paladia” in Malagasy, meaning walking allowance), CHWs often must pay transportation costs out of pocket – or skip visits and activities entirely when they cannot afford it.

> “We CHWs, when we do VAD (visites à domicile; home visits) we go very far, within the fokontany (village). We got phones, bags for TB, we need bicycles because the settlement we go to are very far away.” (male CHW, rural Atsimo-Andrefana)

This need for mobility, as expressed by the CHWs, is directly linked to the lack of financial protection, as outlined in the first dimension.

Provision of personal protective equipment was also cited as essential, particularly when in contact with people with TB. The absence of masks, gloves, or hand sanitizer exposes CHWs to risks and signals neglect compared with better-protected facility-based healthcare workers:

> “We have to buy our own masks to protect ourselves, because when we arrive at 5 a.m. to give the medicine to the patients, the houses are still closed, the air is confined, and that’s a real problem. That’s what made me hesitate to continue this work, but with the knowledge I’ve acquired, I tell myself that if everyone was afraid, the disease would spread even further.” (female CHW, urban Analamanga)

Participants noted that tools and equipment affect how CHWs are perceived: a properly equipped CHW was seen as more competent and credible and gains more respect within the community. In contrast, lacking tools undermined professionalism and weakened authority. Participants emphasized that “equipment” also referred to symbolic or visual items, particularly campaign-specific uniforms, which help signal legitimacy and strengthen CHWs’ visibility during community activities. As one CHW explained:

> “It might be better to adapt the uniforms to the theme of the campaign to give more value to the work of the ACs (CHWs).” (female CHW, urban Analamanga)

### Recognition and integration into the health system

Despite CHWs’ frontline role in the detection, awareness, and patient support, many reported feeling marginalized or seen as lower priority compared to formal healthcare workers:

> “It makes me think a bit and wonder whether the authorities really take ACs (CHWs) seriously. It’s voluntary work, of course, but it’s about time it was taken more seriously.” (female CHW, urban Analamanga)

Participants indicated that this perceived lack of formal recognition had consequences for intrinsic motivation, legitimacy in the community, and collaboration with formal healthcare staff. Several described feelings of discouragement:

> “Sometimes we’re disappointed because, as human beings, it’s true that we do our work for free, but life today is different from what it used to be, so we need to take a closer look at CHWs. And the sad thing is, I admit it, CHWs work hard for the community, yet if one of us dies or falls ill, nobody cares, yet if it’s an artist who dies, he gets a medal. We’ve worked for decades, some have done the job for 40 years, but the state doesn’t consider us.” (female CHW, urban Analamanga).

Concerns were also raised about structural instability of community health programmes, in which CHWs could be dismissed without clear justification:

> “What we insist on is that when the political leadership changes, we should not be pushed aside under the pretext that there are new people. We would not accept that. We are ready to continue as long as possible. It would be unfair to replace us with younger people or those close to local authorities, such as graduates, children of chefs fokontany, or others.” (female CHW, urban Analamanga)

Many CHWs believed that institutional recognition would strengthen credibility, clarify roles, and foster better collaboration with professional healthcare staff. They expressed a desire to be treated as equal partners - with rights of their own - able to report on on-the-ground realities, advocate for community needs, and participate actively in local health decision-making.

## Discussion

CHWs in Madagascar identified five interlinked priorities for effective engagement in the TB response: basic financial protection, standardized and practical training, non-punitive supervision, reliable provision of tools and transport, and recognition within the community and the health system. These priorities were consistent across two contrasting regions.

Despite persistently poor health indicators and minimal progress towards UHC [11] and their essential role in linking the population to essential health services, there is little peer-reviewed evidence capturing CHWs’ experiences and perspectives in Madagascar. Domestic public financing for health remains low; public expenditure on health accounted for only 1.4% of GDP in 2020 [24]. Madagascar therefore receives substantial external assistance, including a US$134.9 million World Bank operation approved in 2022 [25], long-standing Gavi and Global Fund investments [26], and the USAID ACCESS programme (2018-2025) with US$128·5 million [27]. This resonates with broader evidence that fragmented programme architectures weaken standardisation, accountability, and workforce equity, thereby constraining CHW effectiveness [15,28,29]. Furthermore, per diem practices can distort priorities when daily allowances vary across funders, themes, and activities, incentivising CHWs in resource-constrained settings to prioritise higher-pay tasks and partners, a concern consistent with empirical evidence on per diem systems in the health sector, where they can reshape organisational culture, generate, conflict, and change allocation of work time – particularly when rules and rates differ across actors [30,31]. Even when equity and evidence-based principles are applied within isolated projects, these gains often fail to accumulate into a coherent national system, weakening standardisation, accountability, and workforce equity, thereby perpetuating the broader implementation gap. Notably, fragmentation persists despite earlier national efforts to harmonise community health implementation, such as the publication of the Guide de mise en œuvre harmonisée du programme de santé communautaire (Guide MEO) [32] by the Ministry of Health in 2022, which sought to standardise CHW roles, incentives, training, and supervision across partners. Our findings indicate that uneven application and limited accountability for this framework continue to undermine coordination. Addressing this will require strengthened coordination across partners, including aligned incentives, harmonised training and supervision, and adherence to national norms, so that CHWs receive predictable and equitable support.

Our findings also highlight important tensions within the current volunteer-based community health model in Madagascar: while volunteerism remains the formal policy framework, irregular, unpredictable, and fragmented forms of support are framing the reality of CHWs. Concurrent research shows that more than seven out of ten CHW households live on less than US$30 per month [15], with other evidence describing the heavy work demands and livelihood pressures faced by volunteer CHWs, and the tensions this creates for sustained engagement in Madagascar [33]. Comparative evidence from 24 low-income countries clearly demonstrates that remuneration models, whether fixed, performance-based, or hybrid, can enhance motivation, though irregular or unpredictable payments remain a key challenge [34]. Taken together, these findings point to the need for revision of the current national framework and, critically, of how it is implemented by partners.

Importantly, our study shows that CHWs do not uniformly call for full salaried employment. Rather, they emphasise the need for reliable, equitable, and transparent minimum support. In this sense, the core issue is less the volunteer status per se than the absence of predictable material and social protections attached to it. A feasible near-term goal would therefore be to define and finance a nationally standardized minimum CHW support package, including predictable financial incentives or stipends, regular refresher training, essential equipment, transport support, and basic occupational protection.

CHWs also stressed the importance of reliability and fulfilment of promised entitlements by implementing partners and supervisors. This shifts the debate from *what* CHWs need (which is widely known) to *how* accountability can be operationalised so that agreed support is delivered with predictability, equity, and dignity. These findings further highlight a disconnect between CHWs, policy decision-makers, and implementers, sidelining frontline perspectives and contributing to interventions misaligned with on-the-ground realities. This aligns with work emphasising that CHW programme performance is shaped by relational and hierarchical dimensions, including recognition, respect, and the strength of CHWs’ relationships with both communities and the formal health system [28]. Our findings reinforce calls to treat CHWs as rights-holders and system actors, and suggest that reframing CHWs’ role and status serve as a practical governance strategy to improve accountability and reduce the ‘promise–delivery’ gap [35]. In practice, the proposed minimum support package must therefore be complemented by normative standards and accountability requirements for partners, specifying how reliability, transparency, and equity should be upheld, measured, and continuously evaluated in collaboration with CHWs.

Addressing these deficits and establishing an effective national CHW programme will require political will, strengthened governance, and coordination among the many actors involved. While the Malagasy government bears primary responsibility for stewardship, including setting and updating norms, coordinating partners, and monitoring compliance, partners share responsibility for reducing fragmentation by aligning incentives, harmonising training and supervision, sharing operational data, and adhering to a common minimum support package across programmes and geographies. This shared responsibility calls for transparent accountability mechanisms on both sides, including public reporting of partner commitments and delivery against agreed CHW entitlements, and coordination structures with the authority to identify and correct divergences from CHW-oriented norms and standards.

This study provides rare, systematically analyzed qualitative evidence foregrounding CHWs’ perspectives in Madagascar. Its participatory design, inclusion of two contrasting regions, and use of local dialects add depth and transferability. However, the study was limited to focus group discussions in two regions and may not capture the full diversity of CHW experiences nationwide. Using TB as a tracer condition enabled concrete discussions of service delivery realities, particularly those related to stigma, long-term follow-up, infection risk, and referral pathway. At the same time, this foregrounds priorities that are specific to certain infectious diseases and may not be fully generalisable across community health programmes. Nevertheless, the herein identified core system-level enablers are consistent with evidence from other CHW programmes. These limitations do not detract from the central finding of CHWs converging on the same fundamental priorities for effectiveness and equity across contexts.

In conclusion, CHWs in Madagascar remain the cornerstone of community health delivery yet operate frequently without basic means, recognition, and reliability needed to fulfil their role. Addressing these deficits requires coordinated national stewardship and shared accountability among all partners. Strengthening the CHW workforce is not only a matter of fairness but a prerequisite for achieving equitable progress towards UHC in Madagascar.

## Acknowledgments

We thank all those who contributed to this study, including the community health workers and data collectors in the two study districts. We also gratefully acknowledge colleagues at the National Tuberculosis Programme in Madagascar, at both national and regional levels, as well as Doctors for Madagascar, for their support and facilitation.

## Data availability

The data supporting this study are not publicly available due to ethical restrictions. Interested researchers may request access to the data by contacting the corresponding author.

